# Effectiveness of Updated 2023–2024 (Monovalent XBB.1.5) COVID-19 Vaccination Against SARS-CoV-2 Omicron XBB and BA.2.86/JN.1 Lineage Hospitalization and a Comparison of Clinical Severity — IVY Network, 26 Hospitals, October 18, 2023–March 9, 2024

**DOI:** 10.1101/2024.06.04.24308470

**Authors:** Kevin C. Ma, Diya Surie, Adam S. Lauring, Emily T. Martin, Aleda M. Leis, Leigh Papalambros, Manjusha Gaglani, Christie Columbus, Robert L. Gottlieb, Shekhar Ghamande, Ithan D. Peltan, Samuel M. Brown, Adit A. Ginde, Nicholas M. Mohr, Kevin W. Gibbs, David N. Hager, Safa Saeed, Matthew E. Prekker, Michelle Ng Gong, Amira Mohamed, Nicholas J. Johnson, Vasisht Srinivasan, Jay S. Steingrub, Akram Khan, Catherine L. Hough, Abhijit Duggal, Jennifer G. Wilson, Nida Qadir, Steven Y. Chang, Christopher Mallow, Jennie H. Kwon, Bijal Parikh, Matthew C. Exline, Ivana A. Vaughn, Mayur Ramesh, Basmah Safdar, Jarrod Mosier, Estelle S. Harris, Nathan I. Shapiro, Jamie Felzer, Yuwei Zhu, Carlos G. Grijalva, Natasha Halasa, James D. Chappell, Kelsey N. Womack, Jillian P. Rhoads, Adrienne Baughman, Sydney A. Swan, Cassandra A. Johnson, Todd W. Rice, Jonathan D. Casey, Paul W. Blair, Jin H. Han, Sascha Ellington, Nathaniel M. Lewis, Natalie Thornburg, Clinton R. Paden, Lydia J. Atherton, Wesley H. Self, Fatimah S. Dawood, Jennifer DeCuir

## Abstract

**Background:** Assessing COVID-19 vaccine effectiveness (VE) and severity of SARS-CoV-2 variants can inform public health risk assessments and decisions about vaccine composition. BA.2.86 and its descendants, including JN.1 (referred to collectively as “JN lineages”), emerged in late 2023 and exhibited substantial genomic divergence from co-circulating XBB lineages.

**Methods:** We analyzed patients hospitalized with COVID-19–like illness at 26 hospitals in 20 U.S. states admitted October 18, 2023–March 9, 2024. Using a test-negative, case-control design, we estimated the effectiveness of an updated 2023–2024 (Monovalent XBB.1.5) COVID-19 vaccine dose against sequence-confirmed XBB and JN lineage hospitalization using logistic regression. Odds of severe outcomes, including intensive care unit (ICU) admission and invasive mechanical ventilation (IMV) or death, were compared for JN versus XBB lineage hospitalizations using logistic regression.

**Results:** 585 case-patients with XBB lineages, 397 case-patients with JN lineages, and 4,580 control-patients were included. VE in the first 7–89 days after receipt of an updated dose was 54.2% (95% CI = 36.1%–67.1%) against XBB lineage hospitalization and 32.7% (95% CI = 1.9%–53.8%) against JN lineage hospitalization. Odds of ICU admission (adjusted odds ratio [aOR] 0.80; 95% CI = 0.46–1.38) and IMV or death (aOR 0.69; 95% CI = 0.34–1.40) were not significantly different among JN compared to XBB lineage hospitalizations.

**Conclusions:** Updated 2023–2024 COVID-19 vaccination provided protection against both XBB and JN lineage hospitalization, but protection against the latter may be attenuated by immune escape. Clinical severity of JN lineage hospitalizations was not higher relative to XBB lineage hospitalizations.

## Introduction

Estimating COVID-19 vaccine effectiveness (VE) against emerging SARS-CoV-2 variants and describing their clinical severity are important for guiding public health risk assessments and decisions about COVID-19 vaccine composition [1,2]. Updated 2023–2024 COVID-19 vaccines are based on the SARS-CoV-2 Omicron variant XBB.1.5, the predominant lineage circulating in the U.S. during the first half of 2023 [3]. By late 2023, several new variants had emerged, including descendants of XBB lineages and the BA.2.86 variant [4,5].

Initially detected in Denmark and Israel [5], BA.2.86 contains >30 spike mutations compared to XBB.1.5, many of which are associated with increased immune escape and transmissibility [6–12]. The acquisition of the L455S mutation in BA.2.86’s descendant JN.1 was associated with widespread global transmission, and JN.1 became the predominant SARS-CoV-2 lineage in U.S. national genomic surveillance by January 6, 2024 [5]. Descendants of JN.1 have acquired additional spike mutations, including R346T, F456L, and/or T572I, and have increased in prevalence throughout early 2024 [13]. The degree of genomic divergence between BA.2.86 and descendants (hereafter referred to as “JN lineages”) and previously co-circulating XBB lineages, combined with their rapid global spread, raised concern for potential reductions in the effectiveness of updated 2023–2024 COVID-19 vaccines and changes in COVID-19 disease severity.

Early evidence indicated that BA.2.86 and JN.1 infections may be associated with a modest degree of immune escape from updated COVID-19 vaccines [14–20], but were not associated with more severe clinical outcomes compared to previously co-circulating XBB lineages [17,18,21]. Additional estimates of lineage-specific VE and severity are needed given limited evaluations that use whole-genome sequencing in their assessments.

Here, we estimate the effectiveness of an updated 2023-2024 (Monovalent XBB.1.5) COVID-19 vaccine dose (hereafter referred to as an “updated dose”) against XBB and JN lineage hospitalization. We also evaluate relative clinical severity of JN lineage hospitalization compared to XBB lineage hospitalization across a range of in-hospital outcomes.

## Methods

### Design and data collection

This analysis was conducted by the Investigating Respiratory Viruses in the Acutely Ill (IVY) network, a multisite inpatient network comprising 26 hospitals in 20 U.S. states. The IVY network employs a test-negative, case-control design to assess VE; methods have been described previously [22–25]. Briefly, site personnel prospectively enrolled patients aged ≥18 years admitted to IVY network hospitals who met a COVID-19-like illness case definition and received SARS-CoV-2 clinical testing. COVID-19-like illness was defined as ≥1 of the following signs and symptoms: fever, cough, shortness of breath, new or worsening findings on chest imaging consistent with pneumonia, or hypoxemia (defined as an oxygen saturation [SpO2] <92% or supplemental oxygen use for patients without chronic oxygen needs, or escalation of oxygen therapy for patients on chronic supplemental oxygen).

Enrolled patients were tested for SARS-CoV-2 clinically at the local hospital, and additionally had nasal swab specimens shipped to an IVY Network central laboratory at Vanderbilt University Medical Center, where specimens were tested by RT-PCR for SARS-CoV-2, influenza, and respiratory syncytial virus (RSV). In this analysis, case-patients were defined by a positive test result for SARS-CoV-2 in either local or central laboratories for a specimen collected within 10 days of symptom onset and 3 days of hospital admission. Control-patients were defined by a negative result for SARS-CoV-2. Case-patients who tested positive for influenza or RSV were excluded, while control-patients who tested positive for influenza were also excluded due to potential correlation between COVID-19 and influenza vaccination behaviors [26]. Demographic and clinical data were collected through electronic medical record review and patient or proxy interview.

The current analysis included adults admitted to IVY network hospitals from October 18, 2023– March 9, 2024. The analysis period spans the first date on which a patient enrolled in the network was admitted with sequence-confirmed JN lineage infection and the last week during which a patient was admitted with sequence-confirmed XBB lineage infection.

These activities were determined to be public health surveillance with waiver of informed consent by institutional review boards at CDC and each enrolling site and were conducted in accordance with applicable federal law and CDC policy (45 C.F.R. part 46.102(l)(2), 21 C.F.R. part 56; 42 U.S.C. §241(d); 5 U.S.C. §552a; 44 U.S.C. §3501 et seq).

### Classification of vaccination status

Verification of COVID-19 vaccination status was performed using hospital electronic medical records, state vaccination registries, and patient or proxy interviews. Patients were classified into two COVID-19 vaccination groups: 1) those who received an updated 2023–2024 COVID-19 vaccine dose ≥7 days before illness onset, and 2) those who did not receive an updated dose, comprising both patients who had received previous original monovalent and/or bivalent COVID-19 vaccine doses and patients who had never received a COVID-19 vaccine dose. We excluded patients if they received an updated dose <7 days before illness onset, received a non-updated COVID-19 vaccine dose <60 days before their updated dose, received a non-updated vaccine dose after September 10, 2023, received an updated COVID-19 vaccine dose before September 13, 2023, or received >1 updated COVID-19 vaccine dose.

### Severe in-hospital outcomes

Clinical severity of patients with JN and XBB lineage infection was characterized using severe in-hospital outcomes from hospital presentation to the first of hospital discharge, patient death, or hospital day 28 (Supplementary Methods) [23,24]. These outcomes included 1) supplemental oxygen therapy (defined as supplemental oxygen at any flow rate and by any device for those not on chronic oxygen therapy, or with escalation of oxygen therapy for patients receiving chronic oxygen therapy); 2) advanced respiratory support (defined as new receipt of high-flow nasal cannula, non-invasive ventilation, or invasive mechanical ventilation [IMV]); 3) intensive care unit (ICU) admission; or 4) a composite of IMV or death.

### Laboratory methods

Nasal swabs were collected from all enrolled patients and sent to Vanderbilt University Medical Center (Nashville, Tennessee) for RT-PCR testing for SARS-CoV-2, influenza, and RSV (Supplementary Methods). SARS-CoV-2–positive specimens were sent to the University of Michigan (Ann Arbor, Michigan) for whole-genome sequencing to identify SARS-CoV-2 lineages. Sequencing was performed using the Oxford Nanopore Technologies Midnight protocol on a GridION instrument. SARS-CoV-2 lineages were called using PANGO (phylogenetic assignment of named global outbreak lineages) nomenclature (version 4.3.1). Sequences were considered adequate for lineage identification if they had a Nextclade (version 3.4.1) completeness score >80 and a Nextclade quality control overall status of either “good” or “mediocre”. Patients with sequences passing quality control criteria were categorized as either having JN lineage infection/hospitalization (BA.2.86 and its descendants) or XBB lineage infection/hospitalization (all other co-circulating lineages).

### Statistical methods

Descriptive comparisons between COVID-19 case-patients with XBB or JN lineage infections and test-negative control-patients were made using Pearson’s chi-square test for categorical variables and the Kruskal–Wallis test for continuous variables.

### Vaccine effectiveness

We estimated VE against COVID-19–associated hospitalization using multivariable logistic regression to model the odds of case versus control status among patients with and without updated dose receipt. Odds ratios were adjusted for age (18–49, 50–64, ≥65 years), sex (male, female), race/ethnicity (Hispanic or Latino, non-Hispanic Black, non-Hispanic White, non-Hispanic other race, other/unknown), U.S. Health and Human Services (HHS) region, admission date in biweekly intervals, and Charlson comorbidity index (0, 1–2, 3–4, 5–6, ≥7) [27]. VE was calculated as (1 – adjusted odds ratio) × 100% and estimates were calculated separately for case-patients with sequence-confirmed SARS-CoV-2 XBB and JN lineage infections. We conducted sensitivity analyses to evaluate the robustness of findings to different adjustments for potential temporal confounding (Supplementary Results).

### Clinical severity

Multivariable logistic regression was used to estimate the adjusted odds ratio of each severe in-hospital outcome among case-patients with JN lineage hospitalizations compared to those with XBB lineage hospitalizations. Models were adjusted for age (18–49, 50–64, ≥65 years), sex (male, female), race/ethnicity (Hispanic or Latino, non-Hispanic Black, non-Hispanic White, non-Hispanic other race, other/unknown), HHS region, admission date in biweekly intervals, Charlson comorbidity index (0, 1–2, 3–4, 5–6, ≥7), and COVID-19 vaccination status (updated dose, no updated dose). In sensitivity analyses, estimates were stratified by updated dose receipt (in which COVID-19 vaccination status was not adjusted for in regression models) and COVID-19 treatment receipt, defined as receipt of COVID-19 antivirals, steroids, or immunomodulators, for strata with sufficient sample sizes. R (version 4.1.3) was used to conduct all analyses.

## Results

### Description of participants

During October 18, 2023–March 9, 2024, a total of 7,058 patients were admitted with COVID-19–like illness and enrolled in the IVY Network from 26 hospitals in 20 U.S. states. 1,496 (21.2%) patients did not meet inclusion criteria and were removed from the analysis (Supplementary Figure 1). Identification of a SARS-CoV-2 lineage through viral whole-genome sequencing was successful for 62.5% (982/1570) of case-patients meeting all other inclusion criteria. Clinical and demographic characteristics were similar among case-patients with successfully sequenced specimens versus all case-patients (Supplementary Table 1). Among the 5,562 patients meeting all inclusion criteria, 585 (10.5%) were case-patients with sequence-confirmed XBB lineage infection, 397 (7.1%) were case-patients with sequence-confirmed JN lineage infection, and 4,580 (82.3%) were test-negative control-patients.

Among the 5,562 included patients, median age was 67 years (IQR = 55–76 years), 58.9% were non-Hispanic White, 49.9% were male, and 47.9% had a Charlson comorbidity index ≥5 (Table 1). Control-patients had a lower median age (65 years, IQR = 53–75 years) than case-patients with XBB lineage (72 years, IQR = 62–80 years) or JN lineage (71 years, IQR = 60–81 years) infection (*P*<.001); distributions of race/ethnicity and sex were more similar (Table 1). Control-patients had lower median Charlson comorbidity indices (4, IQR = 2–6) than case-patients with XBB lineage (5, IQR = 3–7) or JN lineage (5, IQR = 3–7) infections (*P*<.001). The prevalence of self-reported or documented previous SARS-CoV-2 infection was similar among case-patients with JN lineage infection (27.7%) and control-patients (31.0%), but higher than that among case-patients with XBB lineage infection (16.8%) (*P*<.001). A total of 53 (9.1%) case-patients with XBB lineage infection, 78 (19.6%) case-patients with JN lineage infection, and 844 (18.4%) control-patients had received an updated COVID-19 vaccine dose (Table 1).

**Table 1.**
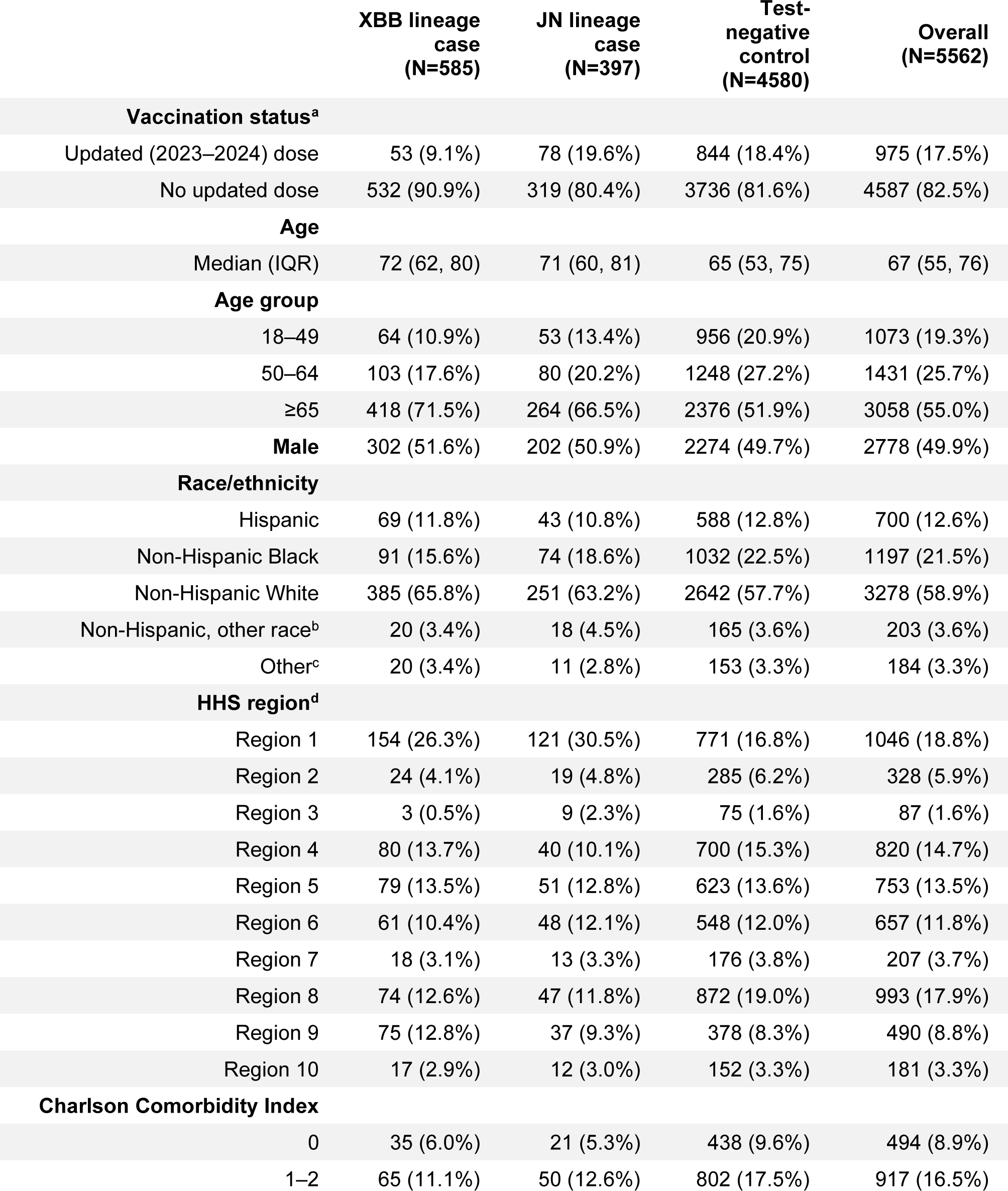
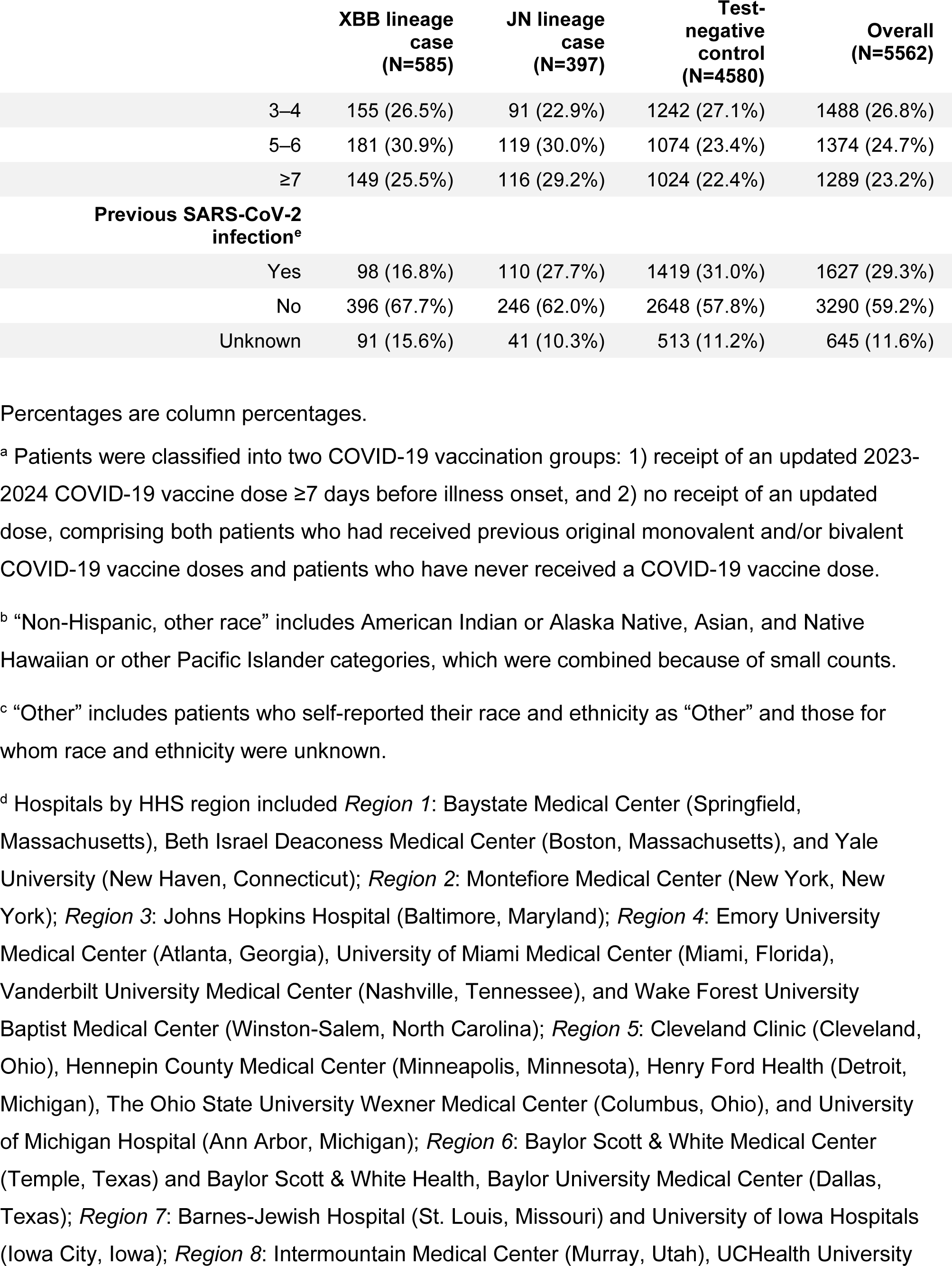
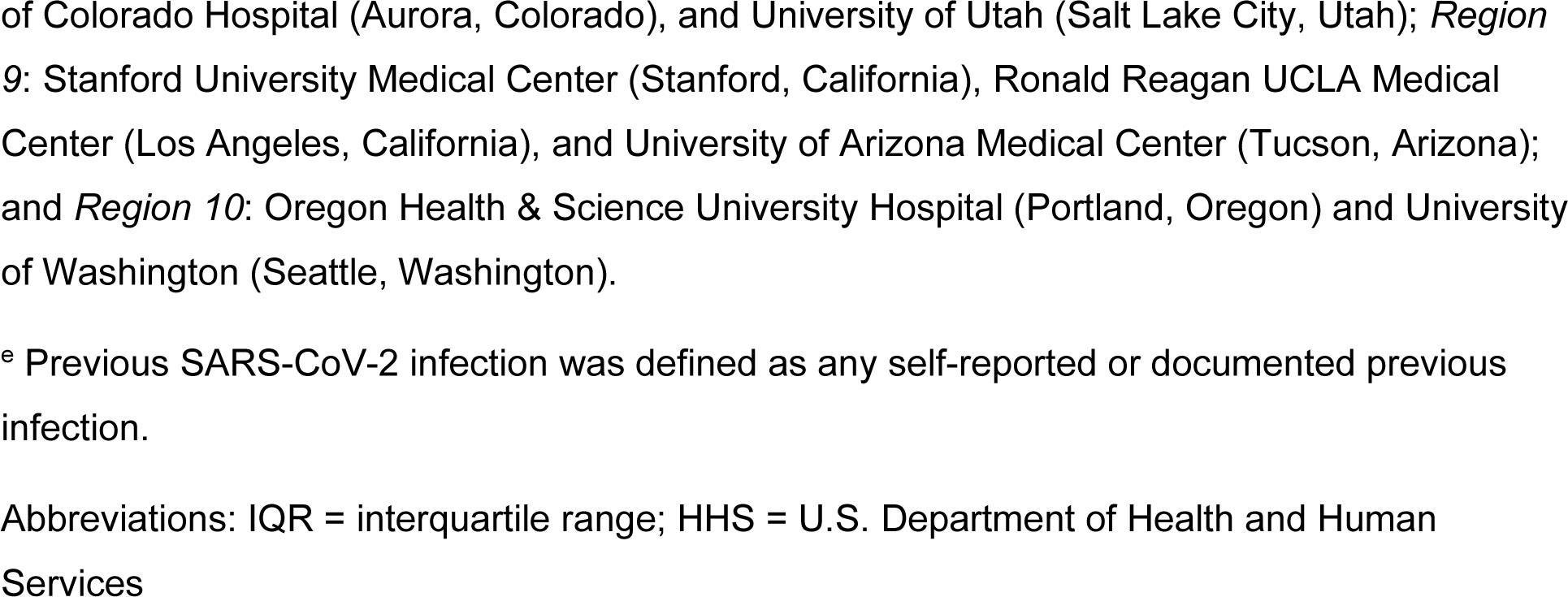
Characteristics of adults hospitalized with COVID-19–like illness by case-control (COVID-19 testing) status and SARS-CoV-2 lineage — IVY Network, 26 Hospitals, October 18, 2023–March 9, 2024.

The most common SARS-CoV-2 lineages among the 585 case-patients with XBB lineage infection were HV.1 (n = 157, 26.8%), FL.1.5.1 (n = 29, 5.0%), and XBB.1.16.6 (n = 26, 4.4%). Among the 397 case-patients with JN lineage infection, 6 (1.5%) were infected with BA.2.86 or BA.2.86.1, 382 (96.2%) with JN.1 or its descendants, and 9 (2.3%) with other descendants of BA.2.86. The proportion of case-patients admitted with JN lineage infection increased steadily beginning in December 2023, and JN lineages became predominant (>50% prevalence) by the week ending January 6, 2024 (Figure 1).

**Figure 1.**
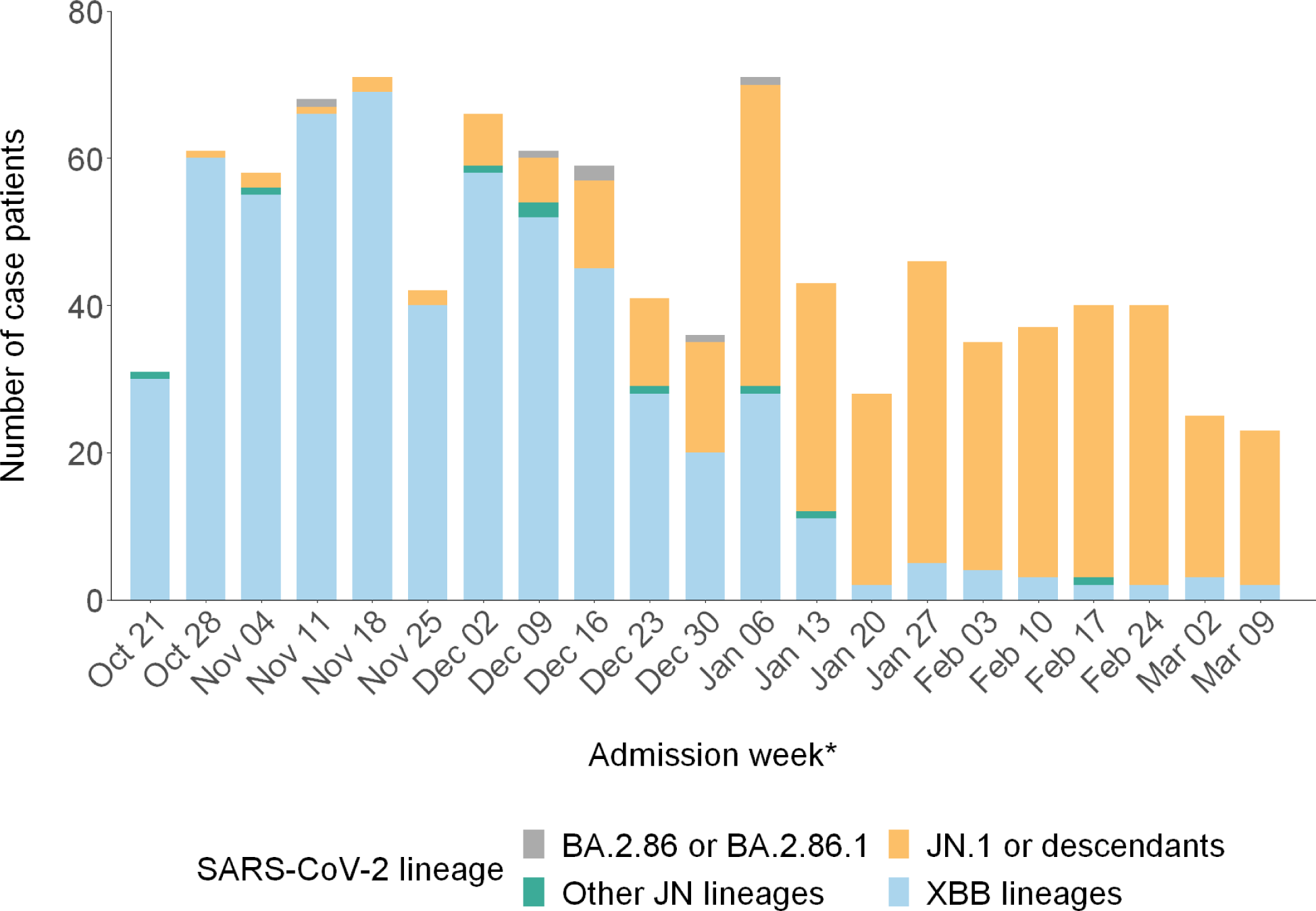
Number of COVID-19 case-patients by hospital admission week and SARS-CoV-2 lineage — IVY Network, 26 Hospitals, October 18, 2023–March 9, 2024. Identification of a SARS-CoV-2 lineage was determined by viral whole-genome sequencing. For analytic purposes, SARS-CoV-2 lineage categories “BA.2.86 or BA.2.86.1,” “JN.1 or descendants,” and “Other JN lineages” were grouped and collectively referred to as “JN lineages.” * Dates are for the end of the admission week.

### Vaccine effectiveness

We estimated the effectiveness of an updated COVID-19 vaccine dose against COVID-19– associated hospitalization due to XBB and JN lineage infection. VE against XBB lineage hospitalization in the first 7–89 days after receiving an updated dose was 54.2% (95% CI = 36.1%–67.1%; median interval since updated dose = 44 days [IQR = 22–67]), and VE against JN lineage hospitalization in the first 7–89 days was 32.7% (95% CI = 1.9%–53.8%; median interval since updated dose = 56 days [IQR = 31–74]) (Figure 2). VE estimates against XBB lineage hospitalization beyond 90 days from vaccination could not be calculated due to the shift in variant predominance from XBB to JN lineages by January 2024. VE against JN lineage hospitalization 90–179 days after receiving an updated dose was 23.4% (95% CI = -11.8% to 47.6%; median interval since updated dose = 118 days [IQR = 107–130]). Estimates were similar when adjusting for time using continuous or categorical calendar time, matching on time using conditional logistic regression, and restricting the analysis period to timeframes during which vaccination coverage was approximately stable (Supplementary Results).

**Figure 2.**
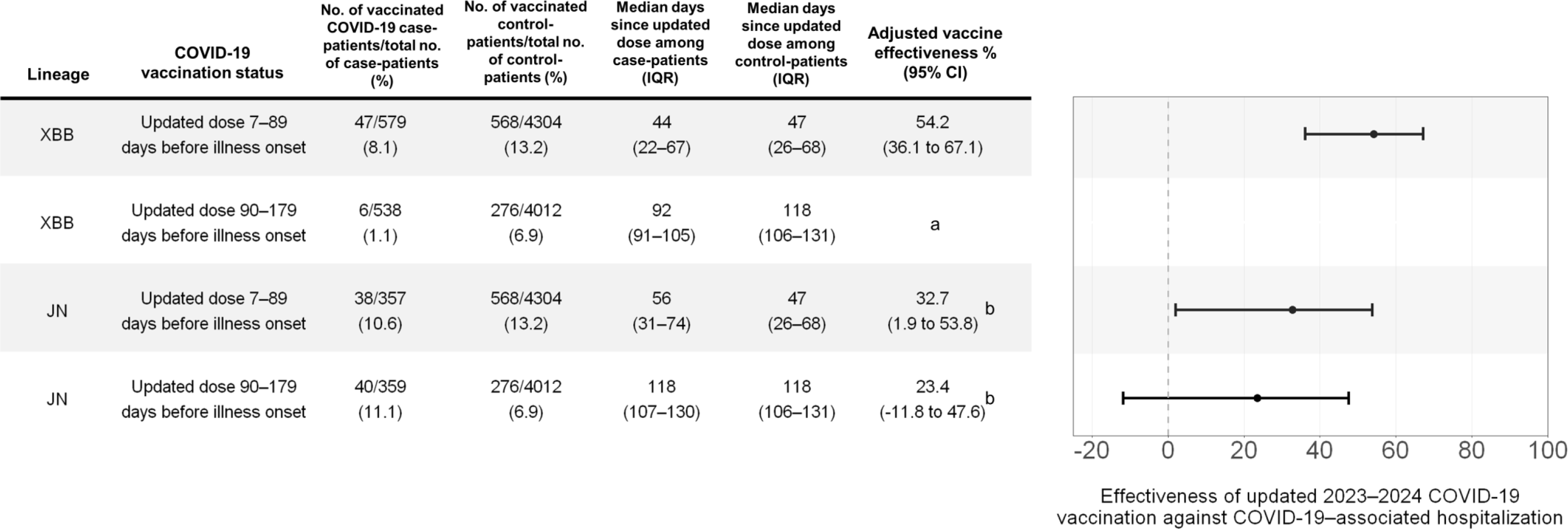
Effectiveness of updated 2023–2024 (monovalent XBB.1.5) COVID-19 vaccination against COVID-19–associated hospitalization from XBB and JN lineage infection — IVY Network, 26 Hospitals, October 18, 2023–March 9, 2024. Vaccine effectiveness was calculated as (1 – adjusted odds ratio) × 100% with odds ratios calculated using multivariable logistic regression adjusting for age, sex, race/ethnicity, HHS region, admission date in biweekly intervals, and Charlson comorbidity index. ^a^ Based on timing of recommendations to receive updated 2023–2024 COVID-19 vaccines and JN lineage emergence, limited numbers of individuals with XBB infection were 90–179 days from their updated dose, precluding estimation of VE within this stratum. ^b^ Some estimates are imprecise, which might be due to a relatively small number of persons in each level of vaccination or case status. This imprecision indicates that the actual vaccine effectiveness could be substantially different from the point estimate shown, and estimates should therefore be interpreted with caution. Abbreviations: IQR = interquartile range; CI = confidence interval; HHS = U.S. Department of Health and Human Services

### Clinical severity

Among 585 case-patients with XBB lineage infections, 366 (62.6%) received supplemental oxygen therapy, 99 (16.9%) received advanced respiratory support, 87 (14.9%) were admitted to an ICU, and 44 (7.5%) received IMV or died. Among 397 case-patients with JN lineage infections, 245 (61.7%) received supplemental oxygen therapy, 76 (19.1%) received advanced respiratory support, 61 (15.4%) were admitted to an ICU, and 32 (8.1%) received IMV or died. The unadjusted proportions of these severe in-hospital outcomes were similar by lineage (Figure 3). The adjusted odds ratios comparing case-patients with JN versus XBB lineage infection for receipt of supplemental oxygen therapy (0.83; 95% CI = 0.54–1.28; *P*=0.40), advanced respiratory support (0.66; 95% CI = 0.38–1.12; *P*=0.12), ICU admission (0.80; 95% CI = 0.46–1.38; *P*=0.42), and IMV or death (0.69; 95% CI = 0.34–1.40; *P*=0.30) indicated no significant differences in clinical severity between lineage groups (Figure 3). Adjusted odds ratios were similar when restricting to patients who received COVID-19 treatment (n = 810, 82.5% of case-patients) and to patients who did not receive an updated vaccine dose (n = 851, 86.7% of case-patients) (Supplementary Figure 4). Estimates were also similar when adjusting for calendar time as a continuous or categorical variable and when matching on time using conditional logistic regression (Supplementary Figure 5).

**Figure 3.**
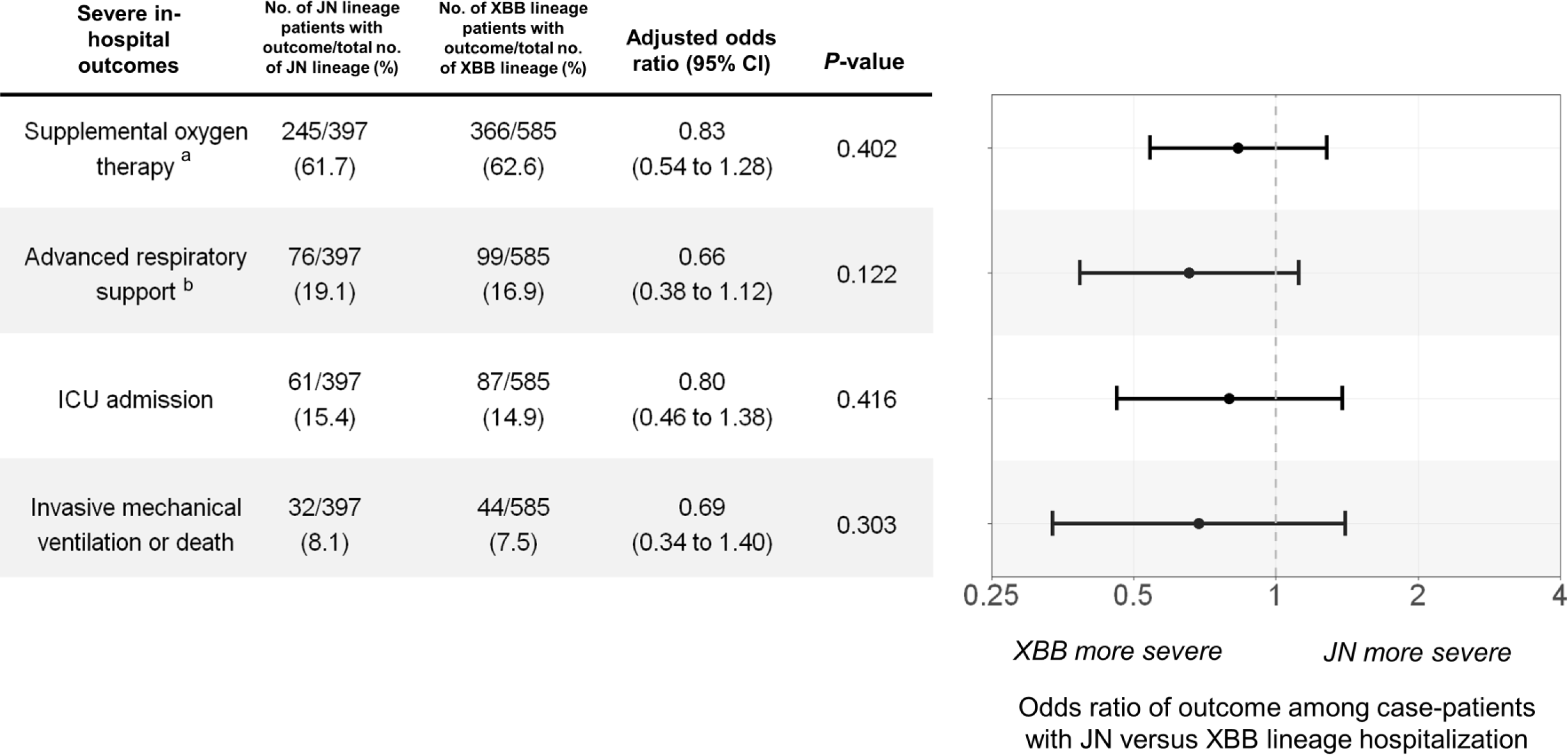
Adjusted odds ratios of severe in-hospital outcomes among adults hospitalized with COVID-19 (JN versus XBB lineage hospitalization) — IVY Network, 26 Hospitals, October 18, 2023–March 9, 2024. Multivariable logistic regression was used to estimate the odds of each outcome among patients with JN versus XBB lineage infection adjusting for age, sex, race/ethnicity, HHS region, admission date in biweekly intervals, Charlson comorbidity index, and COVID-19 vaccination status. ^a^ Supplemental oxygen therapy was defined as supplemental oxygen at any flow rate and by any device for those not on chronic oxygen therapy, or with escalation of oxygen therapy for patients receiving chronic oxygen therapy. ^b^ Advanced respiratory support was defined as new receipt of high-flow nasal cannula, non-invasive ventilation, or invasive mechanical ventilation. Abbreviations: CI = confidence interval; ICU = intensive care unit; IMV = invasive mechanical ventilation

## Discussion

Among adults hospitalized in this multisite surveillance network in the US with whole-genome sequencing-based identification of SARS-CoV-2 lineages, effectiveness of an updated 2023– 2024 COVID-19 vaccine dose in the first 7–89 days was 54% (95% CI = 36%–67%) against XBB lineage hospitalization and 33% (95% CI = 2%–54%) against JN lineage hospitalization, with overlapping confidence intervals between the two lineage groups. These findings indicate that despite substantial genomic divergence of JN lineages from XBB.1.5, updated COVID-19 vaccines continued to provide protection against COVID-19–associated hospitalization. Wide confidence intervals for some estimates precluded confident assessment of the extent to which VE differs by lineage, but point estimates suggest that protection may be lower against JN compared to XBB lineage hospitalization. A lower point estimate for the effectiveness of an updated dose against JN lineage hospitalization 90–179 days after receipt may indicate waning of protection over time, but additional data are needed. We also found that patients hospitalized with JN lineages had similar odds of multiple severe in-hospital outcomes compared to patients hospitalized with XBB lineages, including receipt of supplemental oxygen therapy, advanced respiratory support, ICU admission, and IMV or death.

Although the updated COVID-19 vaccine was effective against JN lineage hospitalization, the lower VE point estimate suggests potential immune escape. This is consistent with emerging evidence from other studies, many of which relied on proxies for determining SARS-CoV-2 lineage, such as S-gene target failure or periods of predominance from national surveillance [13]. In an analysis from the US, VE 60–119 days after dose receipt against symptomatic JN lineage infection determined by S-gene target failure was 49%, 11 percentage points lower than VE against symptomatic XBB lineage infection determined by S-gene target presence [19]. In a period-based analysis using data from the Veterans Affairs healthcare system, VE in the first 60 days against hospitalization during JN.1 predominance was 32%, 30 percentage points lower than VE during XBB predominance [15]. Other approaches have also been used to assess immune escape of JN lineages from COVID-19 vaccines, including a “case-only” study design in which the odds of updated vaccination are compared between patients infected with JN versus XBB lineages. A study using national surveillance data in Denmark estimated an immune escape odds ratio of 1.6 (95% CI = 1.3–2.0), indicating greater immune escape from vaccines for JN lineage infections [17]. Differences in populations, study designs, and median time since updated dose receipt preclude direct comparison of results, but VE point estimates were consistently lower against JN compared to XBB lineages in these studies and in our analysis. These findings are aligned with some immunogenicity data demonstrating slightly increased immune escape of JN.1 compared to XBB descendants, including XBB.1.5, HV.1, and HK.3 [7,12,28].

Our results also expand upon the limited data available on clinical severity of JN lineage infection. Analyses of data from 40 ICUs in France and national surveillance from Denmark found no differences between JN lineages and co-circulating variants on ICU outcomes and ICU/hospitalization risk, respectively [17,21]. A study conducted in an integrated healthcare system in the US found evidence of decreased clinical severity of JN lineage infection for some outcomes, including emergency department visits and hospital admission [18]. The studies available to date and our analysis suggest that JN lineages are unlikely to have *increased* apparent clinical severity compared to XBB lineages, at least in populations with widespread infection-induced and vaccine-induced immunity against SARS-CoV-2 [29].

This analysis is subject to several limitations. First, small sample sizes in some strata due to low uptake of updated COVID-19 vaccines and rare severe in-hospital outcomes limited the precision of some VE and severity estimates, respectively. Second, while we attempted to sequence specimens from all case-patients, we were unable to determine SARS-CoV-2 lineage in a minority of patients due to low viral load and/or viral degradation. Exclusion of this subset of patients did not appear to substantially affect VE estimates (Supplementary Results, Supplementary Figure 6). Third, although IVY is a multisite network with hospitals in geographically diverse locations, our results may not be generalizable to the entire US population. Fourth, analyses did not account for previous SARS-CoV-2 infection, which may provide protection against COVID-19–associated hospitalization and severe outcomes. VE estimates reported in this analysis should therefore be interpreted as the incremental benefit of receiving an updated dose in a population with high levels of infection-induced and/or vaccine-induced immunity [29].

In summary, our results indicate that despite substantial genomic divergence of JN lineages compared with XBB lineages, updated COVID-19 vaccination provided protection against JN lineage hospitalization, although the magnitude of protection may be attenuated [6,7]. Additionally, clinical severity of JN lineage hospitalizations was not higher relative to XBB lineage hospitalizations. In April 2024, the WHO Technical Advisory Group on COVID-19 Vaccine Composition recommended an update to COVID-19 vaccines to include a monovalent JN.1 lineage antigen [2]. Given the potential for continued SARS-CoV-2 evolution, these results highlight the value of estimating lineage-specific VE and clinical severity using active respiratory illness surveillance platforms to guide public health action, including the selection of antigens for use in future COVID-19 vaccines [1,2].

## Data Availability

No additional data available.

## Author contributions

KCM and JD performed data analysis and drafted the manuscript. KCM, JD, DS, FSD, AL, and WHS conceptualized the study design. JD, DS, FSD, and WHS supervised the project. WHS acquired funding. ASL, ETM, AML, and LP conducted whole-genome sequencing. ASL, ETM, AML, LP, MG, CC, RLG, SG, IDP, SMB, AAG, NMM, KWG, DNH, SS, MEP, MNG, AM, NJJ, VS12, JSS, AK, CLH, AD15, JGW16, NQ, SYC, CM, JHK, BP, MCE, IAV, MR, BS, JM, ESH, NIS, JF, YZ, CGG, NH, JDC, KNW, JPR, AB, SAS, CAJ, TWR, JDC, PWB, JHH, and WHS collected the data. All authors contributed to critical review of the manuscript.

## Disclaimer

The findings, conclusions, and views expressed in this presentation are those of the authors and do not necessarily represent the official position of the Centers for Disease Control and Prevention (CDC).

