## Supplementary Materials for "Effectiveness of Updated 2023–2024 (Monovalent XBB.1.5) COVID-19 Vaccination Against SARS-CoV-2 Omicron XBB and BA.2.86/JN.1 Lineage Hospitalization and a Comparison of Clinical Severity — IVY Network, 26 Hospitals, October 18, 2023–March 9, 2024"

INVESTIGATING RESPIRATORY VIRUSES IN THE ACUTELY ILL

The IVY Network

**Contents of Supplementary Materials**

**Supplementary Figure 1.** Participant flow diagram and analytic cohorts ………………………………..10

**Supplementary Table 1.** Characteristics of adults hospitalized with COVID-19 by SARS-CoV-2 sequencing status……………………………………………………………………………………………………….11

Adjusting for confounding by time in vaccine effectiveness and severity analyses……………………..14

**Supplementary Figure 2.** Smoothed change in updated 2023–2024 COVID-19 vaccination coverage by admission week among test-negative controls…………………………………………………….15

**Supplementary Figure 3.** Estimates of updated 2023–2024 COVID-19 vaccine effectiveness against JN lineage hospitalization using different adjustments for confounding by time……………16

**Supplementary Figure 4.** Adjusted odds ratios of severe in-hospital outcomes among adults hospitalized with COVID-19 by COVID-19 vaccination status and receipt of COVID-19 treatment (JN versus XBB lineage hospitalization)………………………………………………………………………………..17

**Supplementary Figure 5.** Adjusted odds ratios of invasive mechanical ventilation (IMV) or death among adults hospitalized with COVID-19 (JN versus XBB lineage hospitalization) using different adjustments for confounding by time……………………………………………………………………………………..19

Assessing biases in selection of specimens to be sequenced and effects on VE…………………..……..…20

**Supplementary Figure 6.** Estimates of updated 2023–2024 COVID-19 vaccine effectiveness among case-patients with SARS-CoV-2 lineage successfully identified through whole-genome sequencing and all case-patients…………………………………………………………………………………………..…21

**Supplementary References**………………………………………………………………………………………………..22

### Appendix A: Investigators and Collaborators

**Investigators and Collaborators in the Investigating Respiratory Viruses in the Acutely Ill (IVY) Network**

**Baylor, Scott & White, Temple, Texas**
Manjusha Gaglani, Tresa McNeal, Shekhar Ghamande Nicole Calhoun, Kempapura Murthy, Joselyn Cravens, Judy Herrick, Amanda McKillop, Eric Hoffman, Ashley Graves, Martha Zayed, Michael Smith

**Baylor, Scott, and White Baylor University Medical Center, Dallas, Texas**
Cristie Columbus, Ashley Bychkowski, Symone Dunkley, Tammy Fisher, Daniela Gonzalez, Robert Gottlieb, Therissa Grefsrud, Mariana Hurutado-Rodriguez, Gabriela Perez

**Baystate Medical Center, Springfield, Massachusetts**
Jay Steingrub, Lesley De Souza, Scott Ouellette

**Beth Israel Medical Center, Boston Massachusetts**
Nathan I. Shapiro, Alessio Barca, Madhavan Das, Jodens Didie, Ana Grafals, Shira Mann, Carlo Ottanelli, Kimberly Redman

**Centers for Disease Control and Prevention (CDC), Atlanta, Georgia**
Kevin Ma, Jennifer DeCuir, Diya Surie, Fatimah Dawood, Katharine Yuengling, Clinton Paden, Lydia Atherton, Natalie Thornburg, Nathaniel Lewis, Sascha Ellington

**Cleveland Clinic, Cleveland, Ohio**
Omar Mehkri, Megan Mitchell, Zachary Griffith, Connery Brennan, Kiran Ashok, Bryan Poynter, Abhijit Duggal

**Emory University, Atlanta, Georgia**
Laurence Busse, Caitlin ten Lohuis, Amy Anderson, Tigist Anemia, Santiago Tovar

**Hennepin County Medical Center, Minneapolis, Minnesota**
Matthew Prekker, Anne Frosch, Audrey Hendrickson, Mary O'Rourke

**Henry Ford Health, Detroit, Michigan**
Ivana A Vaughn, Mayur Ramesh, Lois E Lamerato, Khaled Almawri, Ishraaq Atkins, Jaleesa Clark, Mariia Numi, Shruti Tirumala, Katrina Williams Jean Ashley Lava, Melissa Resk, Sindhuja Koneru, Rachna Jayaprakash, Zina Pinderi

**Intermountain Medical Center, Murray, Utah**
Ithan Peltan, Samuel Brown, Jenna Lumpkin, Shandi Poulsen, Joslyn Bassett, Vineela Thumma

**Johns Hopkins University, Baltimore, Maryland**
David N. Hager, Harith Ali, Minh Phan

**Montefiore Medical Center, Bronx, New York**
Michelle Gong, Amira Mohamed, Rahul Nair, Jen-Ting (Tina) Chen

**Ohio State Medical Center, Columbus, Ohio**
Matthew Exline, Sarah Karow, Maryiam Khan, Connor Snyder, Gabrielle Swoope, David Smith, Brooke Lee, Amanie Rasul, Manisha Pathak, Zachery Lewald, Reece Wilson, Rushil Madan, Jun Sung Park, Rasha Alrifae, Connor Lang

**Oregon Health and Sciences University, Portland, Oregon**
Akram Khan, Catherine L. Hough, Raju Reddy, Shewit P. Giovanni, Aluko A. Hope, Kinsley Hubel, Sherie Gause, Robin Stiller, Bethany Collins, Tomas Cordova

**Stanford University, Stanford, California**
Jennifer G. Wilson, Cynthia Perez, Lily Lau, Ismail Hakki Bekiroglu, Grace Tam, Samantha Ferguson

**University of Arizona**
Jarrod Mosier, Beth Salvagio Campbell, Karen Lutrick

**University of California-Los Angeles, Los Angeles, California**
Nida Qadir, Steven Chang, Cody Tran, Omai Garner, Sukantha Chandrasekaran

**University of Colorado, Aurora, Colorado**
Adit Ginde, Amanda Martinez, Aimee Steinwand, Amy Sullivan, Cori Withers, Jacob Rademacher, Samantha Simon,

**University of Iowa, Iowa City, Iowa**
Nicholas Mohr, Anne Zepeski, Paul Nassar, Noble Briggs, Jacob Hampton, Cathy Fairfield, Heath Gibbs, Courtney Feitsam

**University of Miami, Miami, Florida**
Chris Mallow, Carolina Rivas

**University of Michigan, Ann Arbor, Michigan**
Emily Martin, Arnold Monto, Adam Lauring, Aleda Leis, Leigh Papalambros

**University of Washington, Seattle, Washington**
Nicholas Johnson, Vasisht Srinivasan, Sarah Stucky, Kathyrn Thompson, Joshua Acidera, Katherine Elkort

**Vanderbilt University Medical Center, Nashville, Tennessee**
Wesley H. Self, H. Keipp Talbot, Carlos Grijalva, Ian Jones, Natasha Halasa, James Chappell, Kelsey Womack, Jillian Rhoads, Adrienne Baughman, Colleen Ratcliff, Christy Kampe, Jakea Johnson, Sydney Swan, Cassandra Johnson, Yuwei Zhu, Todd Rice, Jonathan Casey, Yuwei Zhu, Laura L. Short, Lauren J. Ezzell, Margaret E. Whitsett, Rendie E. McHenry, Samarian J. Hargrave, Marcia Blair, Jennifer L. Luther, Claudia Guevara Pulido, Bryan P. M. Peterson, Shanice L. Cummings, Emma Claire Gauthier, Anna C. Jackson, Neekar S. Rashid, Julio Angulo

**Wake Forest University, Winston-Salem, North Carolina**
D. Clark Files, Kevin Gibbs, Leigha Landreth, Lisa Parks, Fay Nketiaa-Agyepong, Darija Ward, Jacqueline Maycee Cain

**Washington University, St. Louis, Missouri**
Jennie Kwon, Bijal Parikh, David McDonald, Carleigh Samuels, Lucy Vogt, Caroline O'Neil, Alyssa Valencia, Francesca Yerbic, Olivia Arter, Kim Vu

**Yale University, New Haven, Connecticut**
Basmah Safdar, Anirudh Goyal, Lauren Delamielleure, Uchechi Okoronkwo, Ivan Velasquez, Carolyn Brokowski, Danielle Paquette

### Appendix B: Supplementary Methods

**1. Severe in-hospital outcomes**

Clinical severity of patients with JN and XBB lineage infection was characterized using the following severe in-hospital outcomes occurring from hospital presentation to hospital discharge, patient death, or hospital day 28:

i) COVID-19-associated supplemental oxygen therapy
ii) COVID-19-associated advanced respiratory support
iii) COVID-19-associated intensive care unit (ICU) admission
iv) COVID-19-associated invasive mechanical ventilation (IMV) or death

<u>i) COVID-19-associated supplemental oxygen therapy</u>

Patients who met the definition of COVID-19-associated supplemental oxygen therapy either required supplemental oxygen therapy at any time during the hospitalization through day 28 for those not on chronic oxygen or, for patients on chronic supplemental oxygen (**Table**), required an escalation in respiratory support. Supplemental oxygen therapy could be delivered at any flow rate and by any device; this included standard flow oxygen (flow rate <30 liters/minute), high-flow nasal cannula (HFNC), non-invasive ventilation (NIV), and IMV. Patients on home IMV prior to the acute illness were not eligible for this outcome.

| Classification of in-hospital respiratory outcome based on type of oxygen or respiratory support used chronically (before illness onset) and highest level received through hospital day 28 | | | | |
|---|---|---|---|---|
| **Chronic pre-illness oxygen use** | **Oxygen use during hospital course (highest support)** | **Is the patient eligible for this outcome?** | | |
| | | **Supplemental oxygen therapy** | **Advanced respiratory support** | **Invasive mechanical ventilation** |
| No oxygen use | Standard flow oxygen | Yes | No | No |
| | High-flow nasal cannula (HFNC) | Yes | Yes | No |
| | NIV | Yes | Yes | No |
| | IMV | Yes | Yes | Yes |
| Standard flow oxygen | Standard flow oxygen | No | No | No |
| | HFNC | Yes | Yes | No |
| | NIV | Yes | Yes | No |
| | IMV | Yes | Yes | Yes |
| Non-invasive mechanical ventilation (NIV) | Standard flow oxygen | No | No | No |
| | HFNC | No | No | No |
| | NIV | No | No | No |
| | IMV | Yes | Yes | Yes |
| Invasive mechanical ventilation (IMV) | Standard flow oxygen | No | No | No |
| | HFNC | No | No | No |
| | NIV | No | No | No |
| | IMV | No | No | No |

ii) COVID-19-associated advanced respiratory support

Patients were classified as having COVID-19-associated advanced respiratory support if they received any of the following during the hospitalization through day 28: HFNC, NIV, or IMV. HFNC was defined as a supplemental oxygen flow rate ≥30 liters per minute. NIV included both continuous positive airway pressure (CPAP) and bilevel positive airway pressure (BiPAP) delivered through a mask. Patients were classified as having NIV use if NIV was received for therapy of the acute illness and not only for treatment of sleep apnea. IMV was defined as positive pressure administered through an endotracheal tube or tracheostomy tube. Patients on home NIV before the acute illness met criteria for the COVID-19-associated advanced respiratory support outcome if they had escalation of respiratory support to IMV in the hospital. Patients on home IMV prior to the acute illness were not eligible for this outcome.

iii) COVID-19-associated intensive care unit (ICU) admission

Patients were classified as having COVID-19-associated ICU admission if they received care in an ICU for any duration of time during the hospitalization through day 28.

iv) COVID-19-associated IMV or death

Patients were classified as having COVID-19-associated IMV or death if they received IMV or died during the hospitalization through day 28. IMV was defined as positive pressure administered through an endotracheal tube or tracheostomy tube. Patients on home IMV prior to the acute illness could not meet the COVID-19-associated IMV or death outcome through receipt of in-hospital IMV.

**2. Laboratory Testing Methods**

At the time of participant enrollment, a nasal swab specimen was collected via a fresh swabbing procedure or collection of a residual aliquot in the clinical laboratory. These specimens were frozen at the enrolling site and shipped to Vanderbilt University Medical Center. Real-time reverse transcription polymerase chain reaction (RT-PCR) testing for SARS-CoV-2, influenza, and RSV was completed at Vanderbilt. Specimens with SARS-CoV-2 detected were then shipped to the University of Michigan for viral whole-genome sequencing. This section describes these laboratory methods.

*SARS-CoV-2 detection by RT-PCR*
Total nucleic acid extract from 100 µl of upper respiratory specimen collected in viral transport medium was prepared using the MagNA Pure LC Total Nucleic Acid Isolation Kit (Roche Molecular Systems, Pleasanton, CA) and MagNA Pure 96 automated extraction platform (Roche) or QiaCube HT automated extraction system (Qiagen, Germantown, MD) and QIAamp 96 Virus QiaCube HT kit (Qiagen). Extracts (100 µl eluate volume) were tested by RT-PCR on the StepOnePlus, QuantStudio 3, or QuantStudio 6 Real-Time PCR System (Applied Biosystems, Waltham, MA) for SARS-CoV-2 nucleocapsid (N)-gene N1 and N2 targets and RNP gene using the CDC protocol, *CDC 2019-Novel Coronavirus (2019-nCoV) Real-Time RT-PCR Diagnostic Panel* with TaqPath 1-Step RT-qPCR Master Mix, CG (Applied Biosystems) (https://www.fda.gov/media/134922/download). The pattern of N1, N2, and RNP Ct values served as a basis to assign a qualitative result of *positive*, *not detected*, *inconclusive*, or *invalid specimen* with respect to SARS-CoV-2 RNA according to interpretive criteria delineated in the assay protocol.

*Influenza detection by RT-PCR*
Total nucleic acid extract from 100 µl of upper respiratory specimen collected in viral transport medium was prepared using the MagNA Pure LC Total Nucleic Acid Isolation Kit and MagNA Pure 96 automated extraction platform or QiaCube HT automated extraction system and QIAamp 96 Virus QiaCube HT kit. Extracts (100 µl eluate volume) were tested by RT-PCR on the StepOnePlus, QuantStudio 3, or QuantStudio 6 Real-Time PCR System for influenza A and B using the CDC Human Influenza Virus Real-Time RT-PCR Diagnostic Panel, Influenza A/B Typing Kit (VER 2) with Superscript III Platinum One-Step Quantitative RT-PCR System containing ROX passive reference dye. Subtyping and lineage identification of influenza A- and B-positive specimens, respectively, by RT-PCR was performed using the CDC Human Influenza Virus Real-Time RT-PCR Diagnostic Panel, Influenza A Subtyping Kit (VER 3) and CDC Human Influenza Virus Real-Time RT-PCR Diagnostic Panel, Influenza B Lineage Genotyping Kit (VER 1.1) with Superscript III Platinum One-Step Quantitative RT-PCR System containing ROX passive reference dye. Each specimen also was tested for RNP using TaqPath 1-Step RT-qPCR Master Mix, CG. PCR reactions consisted of 45 amplification cycles, and Ct values of any magnitude were deemed positive when represented by a characteristic specific amplification curve. A valid influenza A subtype or influenza B lineage identification was contingent on co-detection of the universal influenza type A or B sequence target, respectively. Absence of influenza A and/or B detection in specimens registering RNP Ct values ≥38 was considered inconclusive for the undetected virus(es).

*RSV detection by RT-PCR*
Total nucleic acid extract from 100 µl of upper respiratory specimen collected in viral transport medium was prepared using the MagNA Pure LC Total Nucleic Acid Isolation Kit and MagNA Pure 96 automated extraction platform or QiaCube HT automated extraction system and QIAamp 96 Virus QiaCube HT kit. Extracts (100 µl eluate volume) were tested by RT-PCR on the StepOnePlus, QuantStudio 3, or QuantStudio 6 Real-Time PCR System for a pan-RSV matrix gene target using Superscript III Platinum One-Step Quantitative RT-PCR System containing ROX passive reference dye (Invitrogen, Waltham, MA)

and a screening set of primers and probe (forward: GGCAAATATGGAAACATACGTGAA; reverse: TCTTTTTCTAGGACATTGTAYTGAACAG; probe: FAM-CTGTGTATGTGGAGCCTTCGTGAAGCT-BHQ-1) (Biosearch Technologies, Petaluma, CA). Subgroup differentiation of RSV screen-positive specimens was performed by RT-PCR using Superscript III, a common set of primers, and unique probes targeting A- and B-specific sequences in the viral polymerase (L) gene (forward: AATACAGCCAAATCTAACCAACTTTACA; reverse: GCCAAGGAAGCATGCAATAAA; RSV-A probe: 6FAM-TGCTATTGTGCACTAAAG-MGBNFQ; RSV-B probe: VIC-CACTATTCCTTACTAAAGATGTC-MGBNFQ) (Thermo Fisher, Waltham, MA). Each specimen also was tested for human RNase P (RNP) gene sequence as a marker of specimen adequacy and sensor for PCR inhibitors using TaqPath 1-Step RT-qPCR Master Mix, CG. PCR reactions consisted of 45 amplification cycles, and Ct values of any magnitude were deemed positive when represented by a characteristic specific amplification curve. A valid A or B subgroup identification was contingent on co-detection of the universal RSV target. Absence of RSV detection in specimens registering RNP Ct values ≥40 was considered inconclusive for viral RNA.

*SARS-CoV-2 RNA extraction and whole-genome sequencing*

Specimen aliquots positive by RT-PCR for N1 and N2 targets with a cycle threshold ≤40 at Vanderbilt University Medical Center laboratory were shipped to the University of Michigan on dry ice. RNA was extracted from 200µl transport media with the Thermo Fisher MagMAX Viral Pathogen II Isolation Kit on a KingFisher instrument and eluted in a 50µl volume. Extracted RNA was reverse transcribed with Lunascript RT Supermix (NEB). For each sample, 2 µl of master-mix was added to 8µl of RNA template and incubated at 25°C for 2 min, 55°C for 10 min, 95˚C for 1 min. Viral cDNA was amplified in two multiplex PCR reactions with the Oxford Nanopore Technologies (ONT) Midnight primer pools and protocol using the Q5 Hot Start High-Fidelity DNA Polymerase Master-mix (NEB) with the following thermocycler protocol: 98˚C for 30 s, then 35 cycles of 98˚C for 15 s, 61˚C for 2 min, 65˚C for 3 min. Reaction products for a given sample were pooled together in equal volumes. Sequencing libraries were prepared by adaptor ligation using the ONT Rapid Barcode 96 kit. Three negative control wells (1 HeLa RNA, 2 water) were included on each 96 well RNA harvest plate and carried through the entire process. Barcoded libraries were pooled and sequenced in batches of 96 (GridION instrument). A run was repeated from RNA harvest on if any of the negative controls have >30x read coverage over 10% of the genome. PANGO lineage was assigned on genomes with >80% coverage using Pangolin v4.3.1 (https://pangolin.cog-uk.io, citation in main text). Genomes with >90% coverage were uploaded to GISAID (https://www.gisaid.org/) or NCBI Genbank.

### Appendix C: Supplementary Results

**Supplementary Figure 1.** Participant flow diagram and analytic cohorts

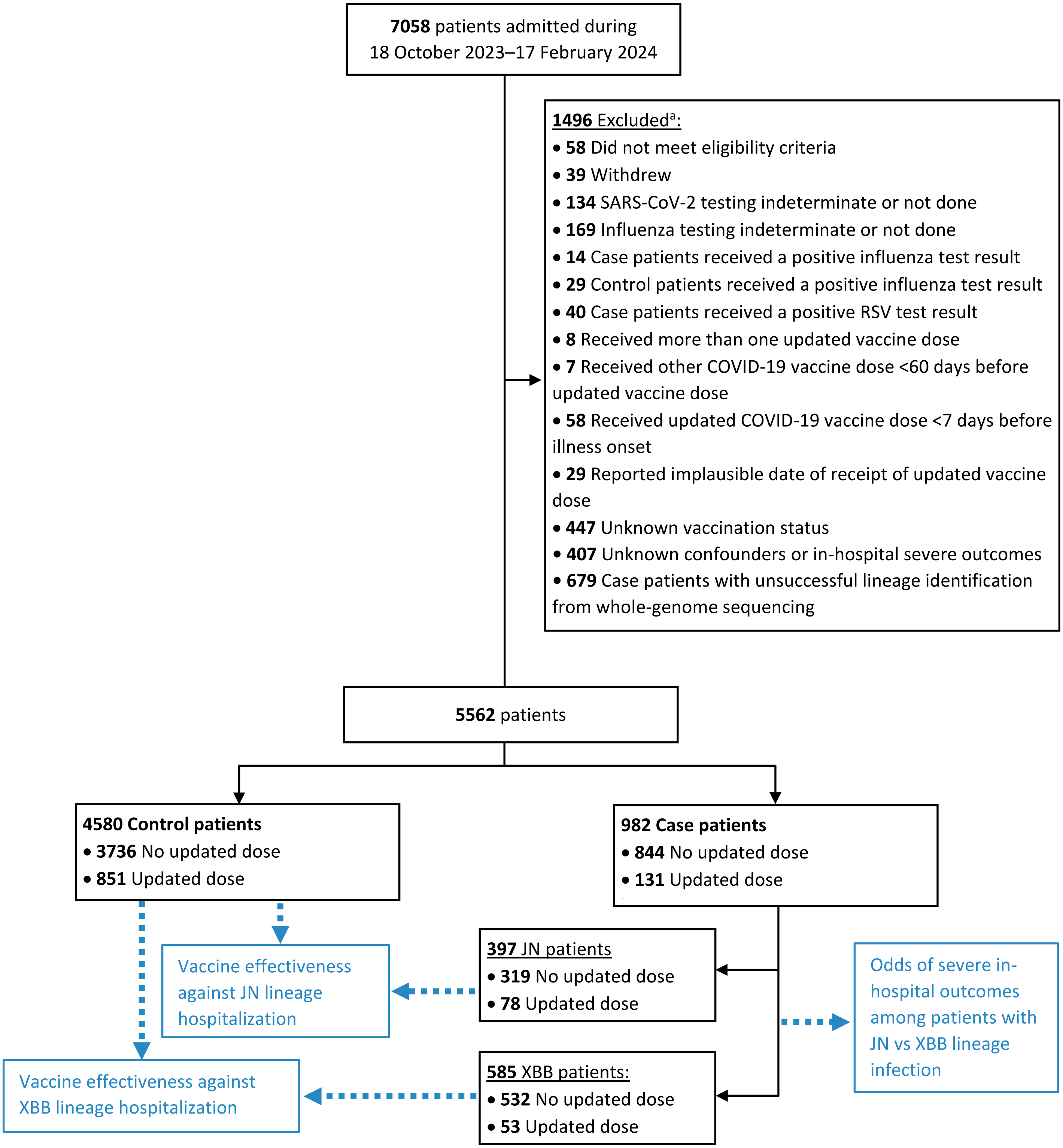

[a] Exclusions not mutually exclusive

**Supplementary Table 1. Characteristics of adults hospitalized with COVID-19 by SARS-CoV-2 sequencing status — IVY Network, 26 Hospitals, October 18, 2023–March 9, 2024.**

| | Sequenced case-patients (N=982) | All case-patients (N=1570) | *P*-value |
|---|---|---|---|
| **Vaccination status[a]** | | | |
| Updated (2023–2024) dose | 131 (13.3%) | 221 (14.1%) | 0.641 |
| No updated dose | 851 (86.7%) | 1349 (85.9%) | |
| **Age** | | | |
| Median (IQR) | 72 (61, 81) | 70 (59, 80) | 0.0151 |
| **Age group** | | | |
| 18–49 | 117 (11.9%) | 216 (13.8%) | 0.0602 |
| 50–64 | 183 (18.6%) | 335 (21.3%) | |
| ≥65 | 682 (69.5%) | 1019 (64.9%) | |
| **Year and biweek** | | | |
| 2023-21 | 31 (3.2%) | 53 (3.4%) | 0.983 |
| 2023-22 | 119 (12.1%) | 176 (11.2%) | |
| 2023-23 | 139 (14.2%) | 209 (13.3%) | |
| 2023-24 | 108 (11.0%) | 175 (11.1%) | |
| 2023-25 | 120 (12.2%) | 188 (12.0%) | |
| 2023-26 | 77 (7.8%) | 117 (7.5%) | |
| 2024-1 | 114 (11.6%) | 179 (11.4%) | |
| 2024-2 | 74 (7.5%) | 131 (8.3%) | |
| 2024-3 | 72 (7.3%) | 127 (8.1%) | |
| 2024-4 | 80 (8.1%) | 124 (7.9%) | |
| 2024-5 | 48 (4.9%) | 91 (5.8%) | |
| **Race/ethnicity** | | | |
| Hispanic | 112 (11.4%) | 185 (11.8%) | 0.897 |
| Non-Hispanic Black | 165 (16.8%) | 286 (18.2%) | |
| Non-Hispanic White | 636 (64.8%) | 995 (63.4%) | |
| Non-Hispanic, other race[b] | 38 (3.9%) | 57 (3.6%) | |
| Other[c] | 31 (3.2%) | 47 (3.0%) | |
| **HHS region[d]** | | | |
| Region 1 | 275 (28.0%) | 395 (25.2%) | 0.645 |
| Region 2 | 43 (4.4%) | 54 (3.4%) | |
| Region 3 | 12 (1.2%) | 27 (1.7%) | |
| Region 4 | 120 (12.2%) | 193 (12.3%) | |
| Region 5 | 130 (13.2%) | 235 (15.0%) | |

| | Sequenced case-patients (N=982) | All case-patients (N=1570) | *P*-value |
|---|---|---|---|
| Region 6 | 109 (11.1%) | 182 (11.6%) | |
| Region 7 | 31 (3.2%) | 52 (3.3%) | |
| Region 8 | 121 (12.3%) | 216 (13.8%) | |
| Region 9 | 112 (11.4%) | 168 (10.7%) | |
| Region 10 | 29 (3.0%) | 48 (3.1%) | |
| **Charlson Comorbidity Index** | | | |
| 0 | 56 (5.7%) | 97 (6.2%) | 0.514 |
| 1–2 | 115 (11.7%) | 219 (13.9%) | |
| 3–4 | 246 (25.1%) | 391 (24.9%) | |
| 5–6 | 300 (30.5%) | 456 (29.0%) | |
| ≥7 | 265 (27.0%) | 407 (25.9%) | |
| **Immunocompromising condition** | | | |
| No | 742 (75.6%) | 1185 (75.5%) | 1 |
| Yes | 240 (24.4%) | 385 (24.5%) | |
| **Previous SARS-CoV-2 infection[e]** | | | |
| Yes | 208 (21.2%) | 379 (24.1%) | 0.156 |
| No | 642 (65.4%) | 970 (61.8%) | |
| Unknown | 132 (13.4%) | 221 (14.1%) | |
| **Supplemental oxygen therapy** | | | |
| No | 371 (37.8%) | 557 (35.5%) | 0.257 |
| Yes | 611 (62.2%) | 1013 (64.5%) | |
| **Advanced respiratory support** | | | |
| No | 807 (82.2%) | 1253 (79.8%) | 0.154 |
| Yes | 175 (17.8%) | 317 (20.2%) | |
| **ICU admission** | | | |
| No | 834 (84.9%) | 1294 (82.4%) | 0.109 |
| Yes | 148 (15.1%) | 276 (17.6%) | |
| **IMV or death** | | | |
| No | 906 (92.3%) | 1433 (91.3%) | 0.422 |
| Yes | 76 (7.7%) | 137 (8.7%) | |

Percentages are column percentages. *P*-values were calculated using Pearson's chi-square test for categorical variables and the Kruskal–Wallis test for continuous variables.

[a] Patients were classified into two COVID-19 vaccination groups: 1) receipt of an updated 2023-2024 COVID-19 vaccine dose ≥7 days before illness onset, and 2) no receipt of an updated dose, comprising

both patients who had received previous original monovalent and/or bivalent COVID-19 vaccine doses and patients who had never received a COVID-19 vaccine dose.

[b] "Non-Hispanic, other race" includes American Indian or Alaska Native, Asian, and Native Hawaiian or other Pacific Islander categories, which were combined because of small counts.

[c] "Other" includes patients who self-reported their race and ethnicity as "Other" and those for whom race and ethnicity were unknown.

[d] Hospitals by HHS region included Region 1: Baystate Medical Center (Springfield, Massachusetts), Beth Israel Deaconess Medical Center (Boston, Massachusetts), and Yale University (New Haven, Connecticut); Region 2: Montefiore Medical Center (New York, New York); Region 3: Johns Hopkins Hospital (Baltimore, Maryland); Region 4: Emory University Medical Center (Atlanta, Georgia), University of Miami Medical Center (Miami, Florida), Vanderbilt University Medical Center (Nashville, Tennessee), and Wake Forest University Baptist Medical Center (Winston-Salem, North Carolina); Region 5: Cleveland Clinic (Cleveland, Ohio), Hennepin County Medical Center (Minneapolis, Minnesota), Henry Ford Health (Detroit, Michigan), The Ohio State University Wexner Medical Center (Columbus, Ohio), and University of Michigan Hospital (Ann Arbor, Michigan); Region 6: Baylor Scott & White Medical Center (Temple, Texas) and Baylor Scott & White Health, Baylor University Medical Center (Dallas, Texas); Region 7: Barnes-Jewish Hospital (St. Louis, Missouri) and University of Iowa Hospitals (Iowa City, Iowa); Region 8: Intermountain Medical Center (Murray, Utah), UCHealth University of Colorado Hospital (Aurora, Colorado), and University of Utah (Salt Lake City, Utah); Region 9: Stanford University Medical Center (Stanford, California), Ronald Reagan UCLA Medical Center (Los Angeles, California), and University of Arizona Medical Center (Tucson, Arizona); and Region 10: Oregon Health & Science University Hospital (Portland, Oregon) and University of Washington (Seattle, Washington).

[e] Previous SARS-CoV-2 infection was defined as any self-reported or documented previous infection.

Abbreviations: IQR = interquartile range; HHS = U.S. Department of Health and Human Services

**Adjusting for confounding by time in vaccine effectiveness and severity analyses**

Confounding by time is a key challenge for vaccine effectiveness (VE) studies particularly when vaccination coverage 1) changes rapidly over time and 2) overlaps with the period when cases of the pathogen of interest are beginning to increase [1,2]. Inclusion of test-negative controls during early analysis weeks when vaccination coverage is low could bias unadjusted vaccine effectiveness estimates downwards, leading to more conservative estimates [1]. Vaccination coverage among test-negative controls reached approximately ~20% by mid-December, which overlapped in time with substantial increases in JN lineage infection beginning in early December (Supplementary Figure 2, Figure 1). Two approaches have been proposed to address temporal confounding in vaccine effectiveness studies: 1) model-based adjustment (either using a conditional logistic model matched on time or by including time as a covariate in an unconditional logistic regression), and 2) analysis period restriction to timeframes when vaccination coverage is approximately stable (December 14, 2023 and onwards) [1,2]; here, we evaluate the sensitivity of our vaccine effectiveness estimates to these adjustments.

We estimated the effectiveness of an updated 2023–2024 COVID-19 vaccine dose in the first 7–89 days after receipt against JN lineage hospitalization (COVID-19–associated hospitalization with JN lineage infection) for the full analysis period (October 18, 2023–March 9, 2024) adjusting for time using different approaches. Vaccine effectiveness in an unadjusted logistic regression model was 21.6% (95% CI = -10.9%–44.7%) (Supplementary Figure 3). Vaccine effectiveness estimates adjusted for demographic covariates (age group, sex, race/ethnicity, HHS region, and Charlson comorbidity index) and for admission date (time) resulted in substantially higher point estimates and confidence intervals excluding the null; estimates were similar when adjusting for categorical week of admission (32.2%, 95% CI = 1.2%–53.5%), categorical biweek of admission (32.7%, 95% CI = 1.9%–53.8%), and admission date modeled using a natural cubic spline with five degrees of freedom (31.9%, 95% CI = 0.4%–53.4%). Estimates were similar using a conditional logistic model matching directly on week (32.1%, 95% CI = 2.0%–52.9%) or biweek (32.6%, 95% CI = 2.8%–53.3%) of admission.

Restriction of the analysis period to timeframes when vaccination coverage is stable is another approach to adjust for temporal confounding that does not require inclusion of time as a covariate in the regression model, but does decrease overall sample sizes. We estimated vaccine effectiveness against JN lineage hospitalization for a shortened analysis period from December 14, 2023–March 9, 2024, coinciding with a period of stable updated vaccination coverage among test-negative controls (Supplementary Figure 2). Vaccine effectiveness was 34.8% (95% CI = 3.5%–56.0%) during this time, which was comparable to the unconditional logistic regression models adjusted for admission week/biweek. Further adjustment for calendar time by including categorical biweek of admission as a covariate did not change estimates, in line with theoretical and simulation results [1,2].

We also evaluated how different adjustments for time affected estimates of severity of JN versus XBB lineage hospitalization. The unadjusted odds ratios comparing case-patients with JN versus XBB lineage infection on the occurrence of invasive mechanical ventilation (IMV) or death was 1.07 (95% CI = 0.67–1.73) (Supplementary Figure 5). Odds ratios adjusted for demographic covariates (age group, sex, race/ethnicity, HHS region, and Charlson comorbidity index) and COVID-19 vaccination status were similar. Additionally adjusting for admission date (time) using categorical biweek of admission shifted point estimates lower (0.69; 95% CI = 0.34–1.40). Estimates were similar when instead adjusting for categorical week of admission, admission date as a linear or natural cubic spline variable, and when using a conditional logistic model matching directly on week or biweek of admission (Supplementary Figure 5).

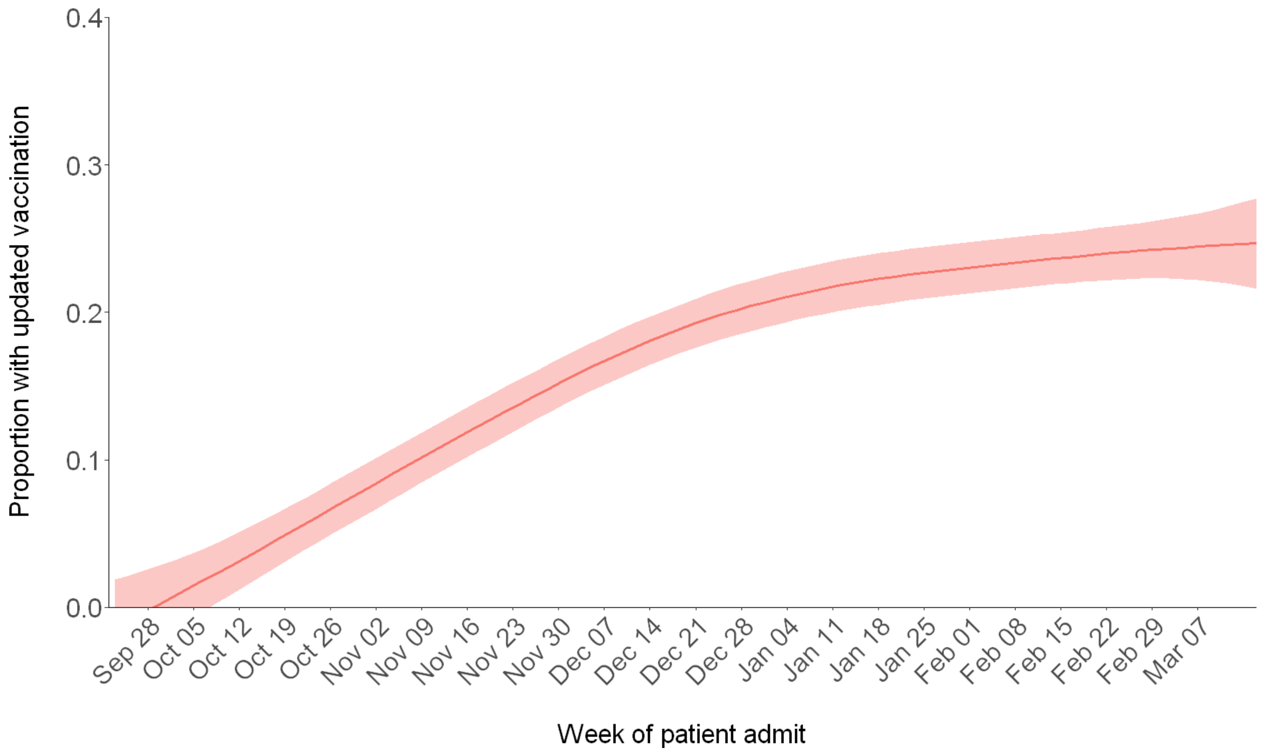

**Supplementary Figure 2.** Smoothed change in updated 2023–2024 COVID-19 vaccination coverage by admission week among test-negative controls. Locally estimated scatterplot smoothing (LOESS) was used to smooth data.

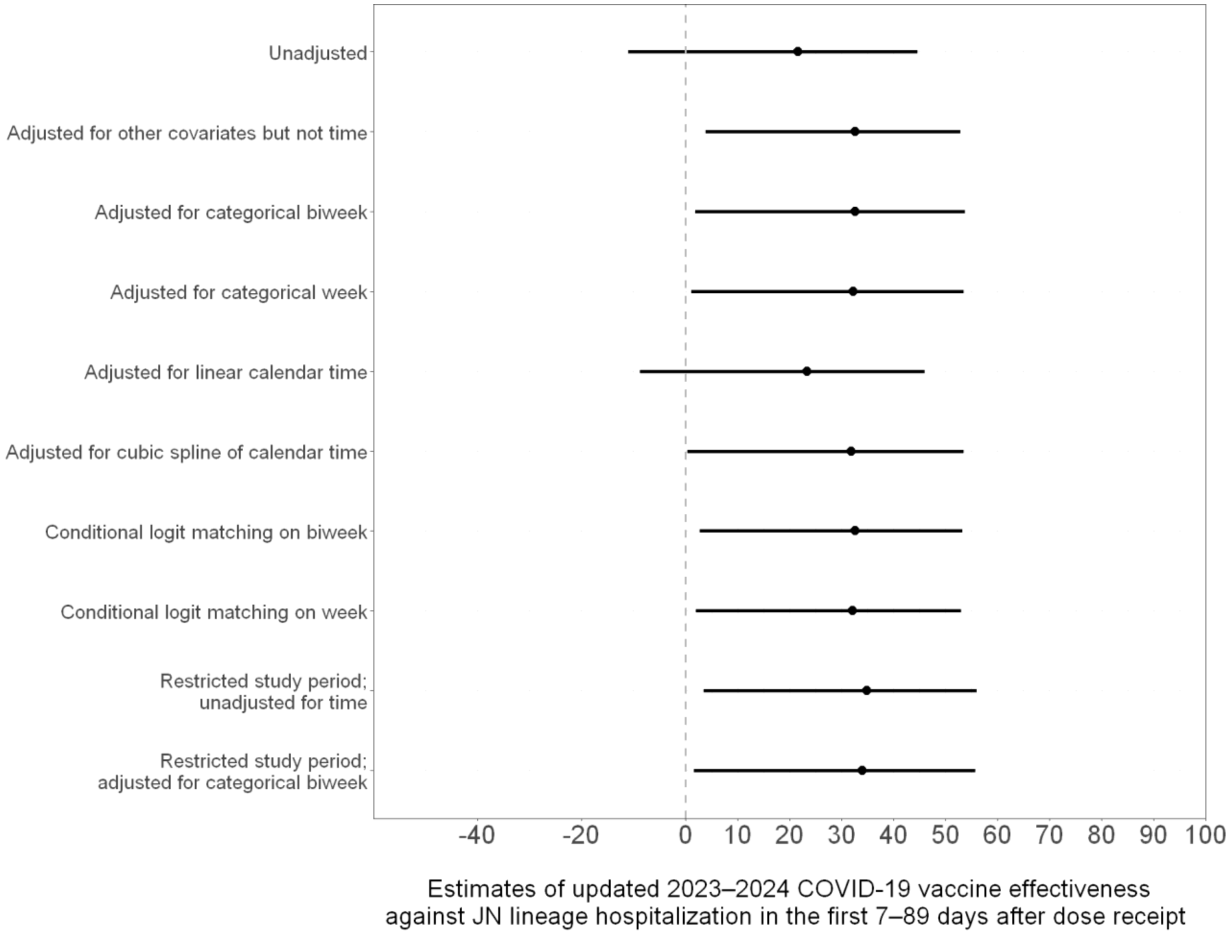

**Supplementary Figure 3.** Estimates of updated 2023–2024 COVID-19 vaccine effectiveness against JN lineage hospitalization in the first 7–89 days after dose receipt using different adjustments for confounding by time.

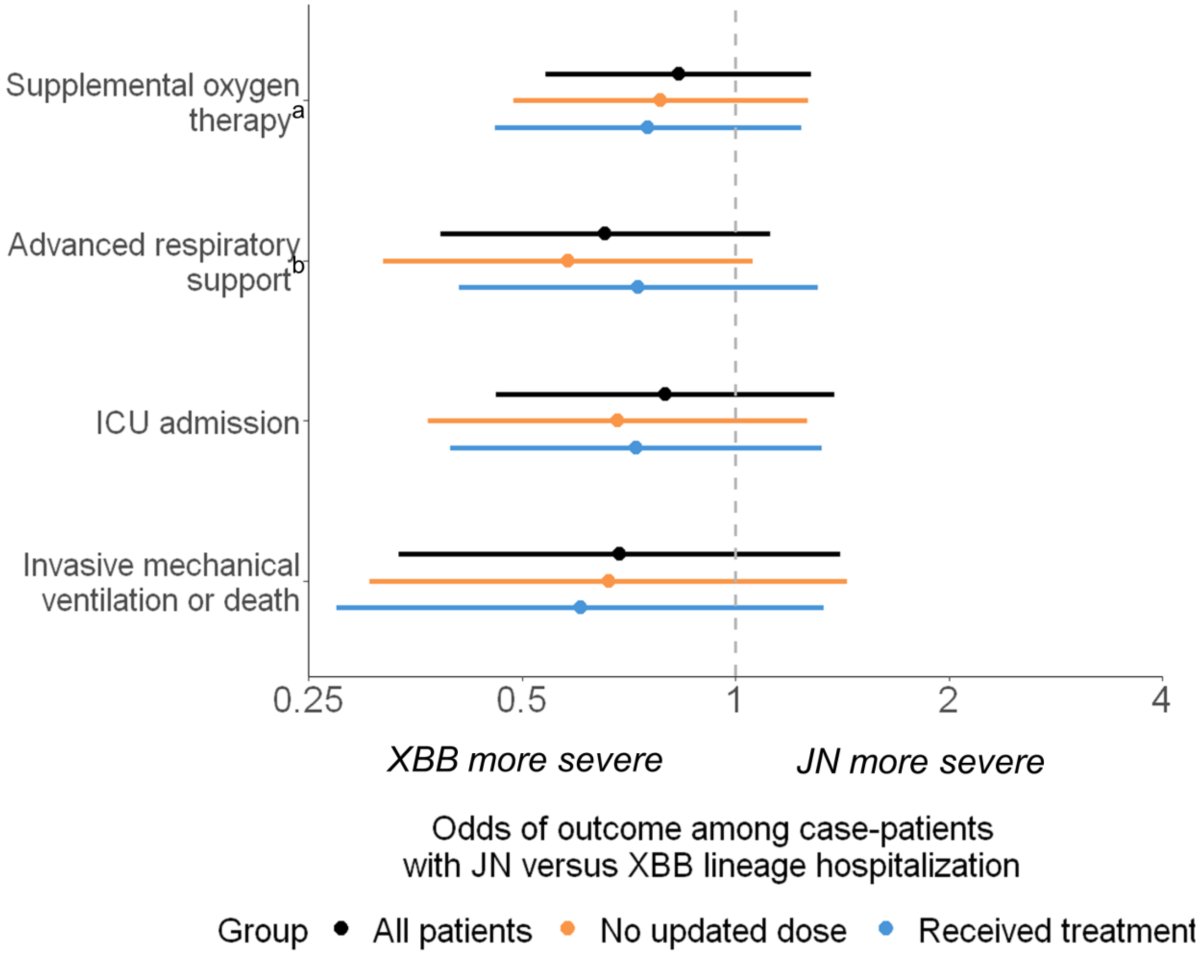

**Supplementary Figure 4.** Adjusted odds ratios of severe in-hospital outcomes among adults hospitalized with COVID-19 (JN versus XBB lineage hospitalization) by COVID-19 vaccination status and receipt of COVID-19 treatment .

Multivariable logistic regression was used to estimate the odds of each outcome among case-patients with JN versus XBB lineage infection, adjusting for age, sex, race/ethnicity, HHS region, admission date in biweekly intervals, Charlson comorbidity index, and COVID-19 vaccination status. Estimates were stratified among patients who did not receive an updated 2023–2024 COVID-19 vaccine dose (n = 851, 86.7% of case-patients) and among patients who received COVID-19 treatment (n = 810, 82.5%), respectively. Limited numbers of patients either received an updated 2023–2024 COVID-19 vaccine dose (n = 131, 13.3%) or did not receive COVID-19 treatment (n = 164, 16.7%), precluding estimation of the odds of severe outcomes within these strata.

Receipt of COVID-19 treatment was defined as receipt of COVID-19 antivirals (outpatient or inpatient remdesivir, Paxlovid, or molnupiravir), steroids (inpatient dexamethasone, methylprednisolone, hydrocortisone, prednisone, or prednisolone), or immunomodulators (inpatient baricitinib, tocilizumab, infliximab, or abatacept). Models stratified by COVID-19 vaccination status did not include COVID-19 vaccination status as a covariate.

[a] Supplemental oxygen therapy was defined as supplemental oxygen at any flow rate and by any device for those not on chronic oxygen therapy, or with escalation of oxygen therapy for patients receiving chronic oxygen therapy.

[b] Advanced respiratory support was defined as new receipt of high-flow nasal cannula, non-invasive ventilation, or invasive mechanical ventilation.

Abbreviations: CI = confidence interval; ICU = intensive care unit; IMV = invasive mechanical ventilation

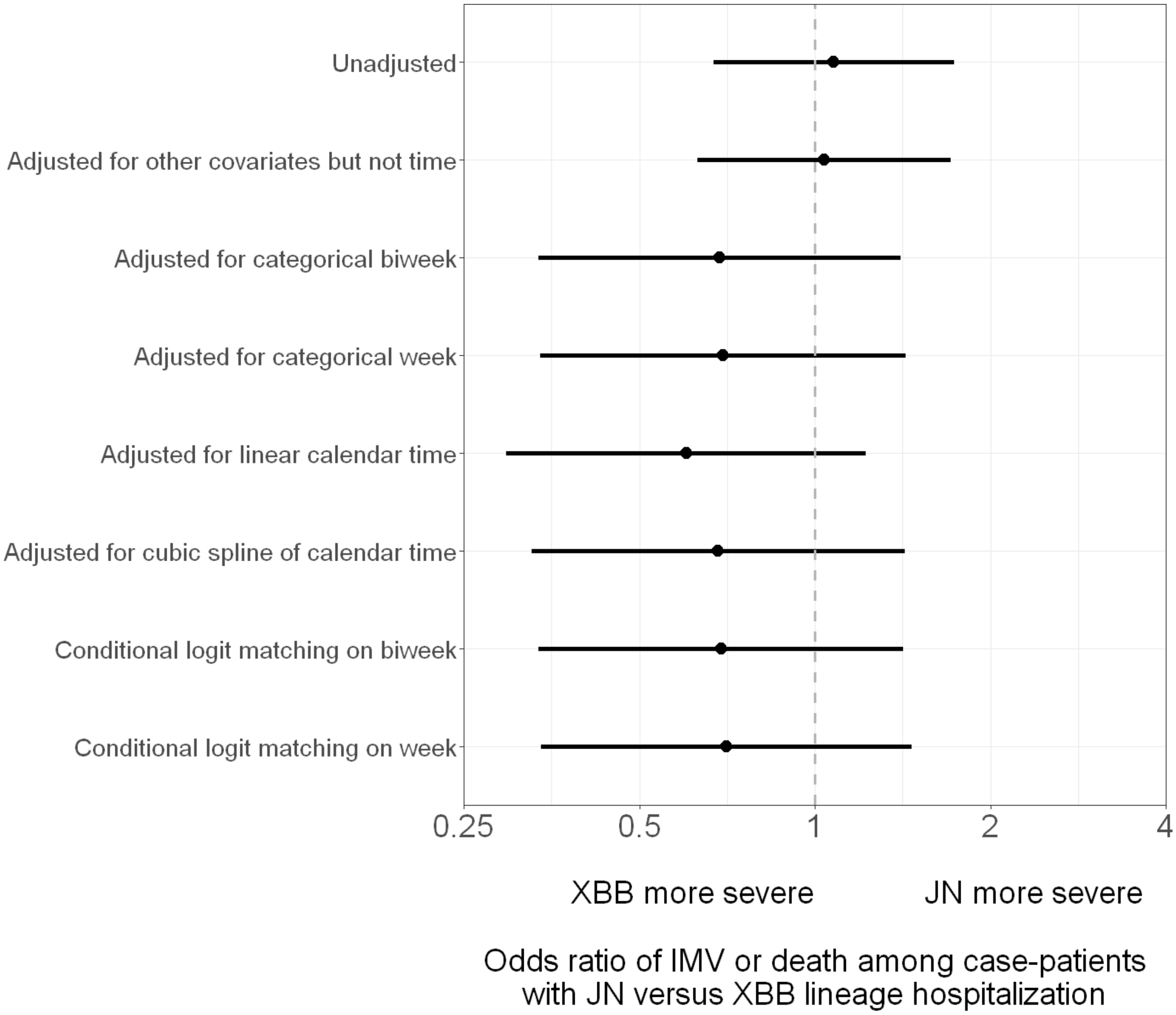

**Supplementary Figure 5.** Adjusted odds ratios of invasive mechanical ventilation (IMV) or death among adults hospitalized with COVID-19 (JN versus XBB lineage hospitalization) using different adjustments for potential confounding by calendar time.

**Assessing biases in selection of specimens to be sequenced and effects on VE**

We assessed how exclusion of case-patients without successful whole-genome sequencing could affect VE estimation. First, we observed that clinical and demographic characteristics were similar among case-patients with successful sequencing versus all case-patients (Supplementary Table 1). Second, per WHO guidance on conducting VE evaluations in the setting of new SARS-CoV-2 variants [3], we estimated VE against COVID-19 hospitalization among all case-patients (regardless of whether lineage identification via sequencing was successful) and compared it to VE among case-patients with successful sequencing results (Supplementary Figure 6). VE against COVID-19 hospitalization overall among all case-patients was 36% (95% CI = 24%–46%) and among sequenced case-patients was 41% (95% CI = 27%–52%), with broadly overlapping confidence intervals. Similar results between all case-patients and sequenced case-patients were observed after stratifying VE by receipt of updated doses 7–89 and 90–179 days earlier. Overall, the comparable estimates suggested that exclusion of the minority of patients without successful sequencing did not result in biases when estimating VE.

| Patient category | COVID-19 vaccination status | No. of vaccinated COVID-19 case-patients/total no. of case-patients (%) | No. of vaccinated control-patients/total no. of control-patients (%) | Median days since updated dose among case-patients (IQR) | Median days since updated dose among control-patients (IQR) | Adjusted vaccine effectiveness % (95% CI) |
|---|---|---|---|---|---|---|
| Sequenced | Any updated dose | 131/982 (13.3) | 844/4580 (18.4) | 70 (38–106) | 68 (36–105) | 40.5 (26.6 to 51.7) |
| Sequenced | Updated dose 7–89 days earlier | 85/936 (9.1) | 568/4304 (13.2) | 52 (25–68) | 47 (26–68) | 46.4 (31.3 to 58.3) |
| Sequenced | Updated dose 90–179 days earlier | 46/897 (5.1) | 276/4012 (6.9) | 114 (104–126) | 118 (106–131) | 22 (-10.7 to 45.1) |
| All | Any updated dose | 221/1570 (14.1) | 844/4580 (18.4) | 68 (37–100) | 68 (36–105) | 35.6 (23.8 to 45.7) |
| All | Updated dose 7–89 days earlier | 148/1497 (9.9) | 568/4304 (13.2) | 48 (24–68) | 47 (26–68) | 38.9 (25.5 to 50) |
| All | Updated dose 90–179 days earlier | 73/1422 (5.1) | 276/4012 (6.9) | 111 (100–126) | 118 (106–131) | 24.3 (-0.9 to 43.3) |

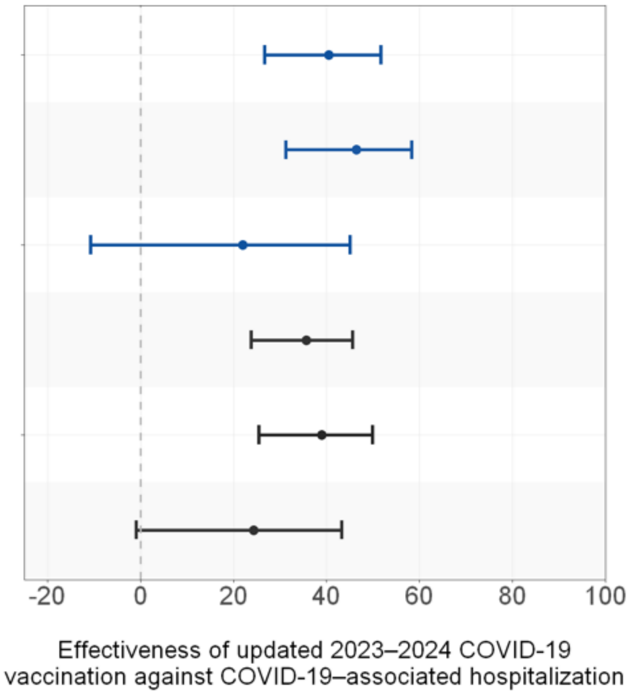

**Supplementary Figure 6.** Estimates of updated 2023–2024 COVID-19 vaccine effectiveness among case-patients with SARS-CoV-2 lineage successfully identified through whole-genome sequencing and among all case-patients. Vaccine effectiveness was calculated as (1 – adjusted odds ratio) × 100% with odds ratios calculated using multivariable logistic regression adjusting for age, sex, race/ethnicity, HHS region, admission date in biweekly intervals, and Charlson comorbidity index.

Abbreviations: IQR = interquartile range; CI = confidence interval; HHS = U.S. Department of Health and Human Services
